# Sex differentiated and domain specific patterns of longitudinal cognitive decline across subjective cognitive decline and amyloid positivity groups

**DOI:** 10.64898/2026.05.06.26352563

**Authors:** Cassandra Morrison, Mahsa Dadar, Yashar Zeighami

## Abstract

**Background:** Subjective cognitive decline (SCD) is associated with increased cognitive impairment and dementia. However, limited research has explored how amyloid (A) pathology contributes to these cognitive changes over time and whether these changes differ by sex.

**Methods:** 1185 cognitively normal older adults (955 A-, 230 A+; 959 SCD-, 226 SCD+) from the National Alzheimer’s Coordinating Center dataset were included. Linear mixed effects examined the interactions between SCD, sex, and amyloid positivity in predicting cognitive decline.

**Results:** SCD+ and A+ individuals exhibited increased global cognition declines (*p*<.05), and A+SCD+ individuals showed the steepest decline in global cognition and function status (*p*<.05). A+ males exhibited increased functional deficits (*p*<.05), while A+SCD+ females exhibited increased language deficits (*p*<.05).

**Discussion:** Our findings suggest that SCD and amyloid-positivity differentially impact global cognition, functional status, and language in males versus females, with important implications for clinical trials and therapeutic interventions.

**Highlights:**

- Few studies have explored the independent and joint effects of amyloid and sex in SCD
- SCD is associated with increased rates of global cognitive decline
- Amyloid positive females with SCD exhibit increased language declines
- Amyloid positive males exhibit increased functional status declines

**Research in Context:** *Systematic review:* We reviewed the literature using traditional sources (e.g., PsycInfo, PubMed) and found that there are limited findings exploring longitudinal cognitive trajectories in people who are amyloid positive with SCD and whether these trajectories differ by sex.

*Interpretation:* Our findings suggest that SCD and amyloid positivity jointly interact to influence global cognitive and functional declines. Females experience language deficits when they have both SCD and amyloid positivity whereas males with amyloid positivity exhibit increased functional deficits. Together these findings suggest that SCD status and amyloid positivity differentially impact females and males.

*Future directions:* More research is needed using grouping amyloid, tau, neurodegeneration, and vascular pathologies together to explore the joint impact on cognitive change and conversion in people with SCD.

## Introduction

Subjective cognitive decline (SCD) is an early indicator of cognitive decline, representing a potential preclinical stage of dementia^1–3^. SCD is identified based on self-reported experiences of cognitive deficits such as concerns about memory loss in the absence of objectively measurable cognitive deficits^1–3^. Older adults with SCD typically exhibit increased decline in global cognition, memory, language, and executive functioning compared to those without SCD^4–6^. However, these results are inconsistent in cross-sectional studies (see Zhou et al., 2024 for review)^7^. Complementing the increased rates of decline, people with SCD also experience higher rates of dementia-related brain changes compared to those without SCD^8–13^. These cognitive and brain changes observed in SCD align with findings that older adults experiencing SCD are at a higher risk of developing mild cognitive impairment (MCI) and dementia compared to those without SCD^2,14–16^.

The International Subjective Cognitive Decline Initiative (SCD-I) Working Group has developed a conceptual framework for research on SCD as a preclinical feature of Alzheimer’s disease (AD)^1^. This framework includes SCD *plus*, a category that increases risk of conversion to AD in those with SCD. Features that increase the likelihood include persistent feelings of subjective decline in memory, onset within the last five years and after the age of 60, worry about SCD, feelings of worse performance than others, seeking of medical help, confirmation of declines by an informant, presence of APOE ε4 genotype, and biomarker evidence for AD^1,2^. AD-related biomarker evidence includes presence of amyloid β and tau protein pathology, which develop before clinical symptoms, representing asymptomatic AD (i.e., preclinical AD)^2^.

Currently, new AD treatments such as lecanemab^17^ and donanemab^18^ target amyloid to reduce the buildup of plaques^19^. Amyloid has been a major therapeutic target because its accumulation occurs early in AD pathogenesis^20^, can be detected years before symptoms onset using biomarkers such as PET and CSF measures^20^, and is hypothesized to facilitate tau pathology and neurodegeneration^21^. Targeting amyloid early in the disease process may thus help mitigate downstream pathological changes and delay cognitive decline. Given that amyloid deposition occurs years before a diagnosis^20^, research must explore its role early, during the preclinical stages of dementia, including SCD. Some studies report greater cognitive decline among amyloid-positive compared to amyloid-negative individuals^22–24^, however, this group difference has not always been observed^25^. Additionally, one study found that amyloid positivity predicted cognitive decline in those with SCD^26^.

The relationships between amyloid, SCD, and cognitive decline may also be influenced by sex. Among individuals with SCD, females are reported to show increased rates of cognitive decline^6^ and to receive a future dementia diagnosis compared to males^27^. Females are also disproportionately affected by AD, with prior research indicating they are approximately twice as likely than males to develop AD^28–30^. Among those with AD, females exhibit more pathological burden^31^ and faster cognitive deterioration^32,33^ than their male counterparts. Furthermore, evidence suggests that lecanemab, an amyloid-targeting drug, may be less effective in females than males^34^. This is consistent with prior findings showing that, among amyloid-positive tau-negative individuals, males exhibit faster progression than females^35^. Together, these findings suggest that examining sex differences in amyloid and SCD interactions may clarify whether amyloid pathology is associated with different cognitive trajectories in SCD.

While some studies have examined the interaction between amyloid positivity and SCD^22–26^, these studies are limited by small samples, short follow-up durations, limited exploration of various cognitive domains, and no examination of sex differences. More research is needed to improve our understanding of how amyloid pathology influences cognitive decline trajectories in SCD. The current study leverages the National Alzheimer’s Coordinating Center (NACC) dataset with extensive follow-up times to investigate the impact of amyloid on cognitive and functional change in cognitively healthy older adults with SCD and whether these changes differ by sex.

## Methods

Measurements were obtained from the NACC database (https://naccdata.org/). All participants provided informed consent and the necessary ethics approvals were obtained by the local institutional review board from each NIA Alzheimer’s Disease Research Center (ADRC) that contributed the data to NACC. As described previously, PET and CSF based binary indicators of elevated amyloid and tau were obtained from the NACC database, provided by the NACC ADRCs (https://files.alz.washington.edu/documentation/uds3-tip-ded.pdf)^35^. Participants aged above 55 and under 95 and with no diagnoses of neurological disorders at baseline (based on NACCETPR variable) were included if they had longitudinal cognitive assessments as well as baseline demographics information and SCD assessments. Similar to our previous work^35^, PET or CSF based measures of amyloid positivity provided by the NACC were used to determine amyloid classifications (A+, Amyloid Positive; A-Amyloid Negative).

Functional impairments were assessed using the Functional Activities Questionnaire (FAQ) questionnaire (N = 1,185; 7,443 longitudinal visits). Global cognition was examined using the Clinical Dementia Rating Sum of Boxes (CDR-SB) scale (N = 1,185; 7,443 visits). Executive functioning was assessed using Trail Making Test (TMT) Parts A (N = 1,162; 6,237 visits) and B (N = 1,160; 6,221 visits). Semantic functioning was assessed using Animals (N = 1,182; 7,035 visits) and Vegetables (N = 1,180; 7,023 visits) tests. The estimates for Animals and Vegetables were multiplied by (–1) so that positive estimates consistently indicate worse cognitive performance for all tests.

### Statistical Analyses

Group differences at baseline were assessed using two-way analysis of variance (ANOVA), with SCD status (SCD− vs SCD+) and amyloid status (A− vs A+) as between-subject factors, including their interaction term. To examine group differences in the proportion, we fitted a generalized linear model with binomial distribution and logit link function. SCD status, amyloid status, and their interaction were included as predictors.

Mixed effects models were used to examine if the trajectories of longitudinal cognitive decline differed between SCD+ versus SCD-.

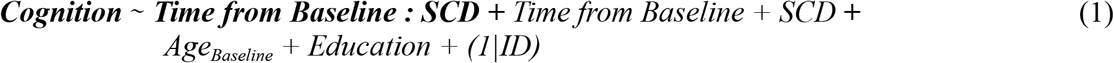

*Cognition* was included as the dependent variable indicating performance on different cognitive tests (i.e., CDR-SB, FAQ, Trail A and B, and Animals and Vegetables scores). *Time from Baseline* indicates the time between each longitudinal timepoint and the baseline clinical assessment visit. SCD is a categorical variable contrasting SCD+ participants against SCD-participants. *Age*_*Baseline*_ and *Education* were included as fixed covariates in all models. The variables of interest were ***Time from Baseline***, reflecting change in cognition over time in the reference group (e.g. SCD-), ***SCD***, reflecting baseline differences in cognition between groups, and ***Time from Baseline : SCD***, reflecting potential differences in rates of cognitive decline across SCD groups.

Similarly, longitudinal mixed effects models were used to examine the differences in rates of cognitive decline of SCD+ versus SCD-participants across amyloid positivity groups:

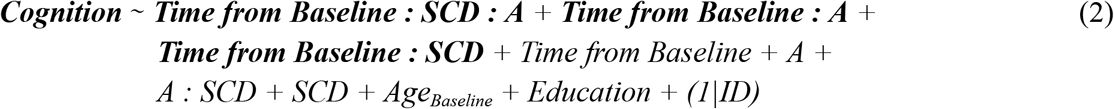

Where *A* indicates a categorical variable reflecting amyloid positivity status. The variable of interest was ***Time from Baseline : A : SCD, Time from Baseline : A***, and ***Time from Baseline : SCD***, reflecting potential differences in rates of cognitive decline across SCD and amyloid groups.

Finally, an interaction with *Sex* was added to the model to investigate potential sex differences in the cognitive decline trajectories:

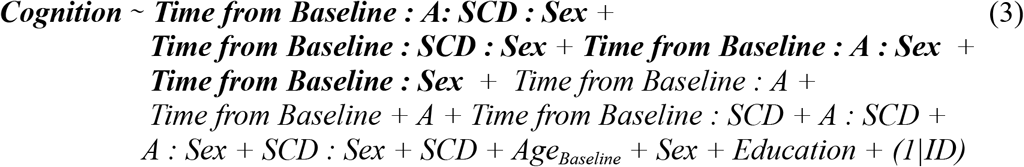

In these models, the variables of interest were ***Time from Baseline : A: SCD : Sex, Time from Baseline : SCD : Sex, Time from Baseline : A : Sex***, and ***Time from Baseline : Sex***, reflecting potential sex differences in rates of cognitive decline across SCD and amyloid groups.

The differences in the proportion of individuals that experience cognitive decline, defined as drop in CDR-SB scores between the first and last available visits, were compared across groups of interest using *χ*2 tests. The results were corrected for multiple comparisons using the false discovery rate (FDR) controlling method^36^.

## Results

Table 1 summarizes the baseline characteristics of the participants included in this study. No significant differences were observed between groups in any of the variables except for age at baseline (p < 0.001). Post hoc comparison revealed no significant differences in age at baseline between SCD+ and SCD-groups (p > 0.2), while A+ participants were significantly older than A-participants (A+: 70.6 ± 6.92, A-: 67.8 ± 6.62, p < 0.001). Furthermore, APOE ε4 positivity differed significantly by amyloid status (OR = 2.30, p < .001), with no main effect of SCD status (p = .66) and a trend-level SCD × amyloid interaction (p = .065).

**Table 1.** Baseline characteristics of the participants included in this study.

| Groups | SCD- |  |  |  | SCD+ |  |  |  |
| --- | --- | --- | --- | --- | --- | --- | --- | --- |
|  | Females |  | Males |  | Females |  | Males |  |
| Variables | Amyloid - | Amyloid + | Amyloid - | Amyloid + | Amyloid - | Amyloid + | Amyloid - | Amyloid + |
| N | 503 | 101 | 285 | 70 | 119 | 38 | 48 | 21 |
| Age | 67.1 (6.4) | 70.3 (6.4) | 68.6 (6.7) | 71.7 (7.3) | 68.4 (7.0) | 69.9 (7.0) | 68.7 (6.9) | 69.7 (8.0) |
| APOE<sup>+</sup> | 120 (27.0%) | 35 (40.2%) | 73 (29.0%) | 28 (43.8%) | 22 (21.6%) | 17 (50.0%) | 14 (35.0%) | 13 (65.0%) |
| Education | 16.5 (2.3) | 16.2 (2.3) | 17.0 (2.3) | 17.2 (2.4) | 16.0 (2.4) | 16.7 (2.4) | 17.2 (2.1) | 17.3 (2.5) |
| FAQ | 0.05 (0.34) | 0.06 (0.31) | 0.07 (0.44) | 0.03 (0.17) | 0.15 (0.53) | 0.11 (0.39) | 0.21 (0.68) | 0.24 (0.77) |
| Trail A | 28.5 (10.7) | 28.5 (8.7) | 28.1 (9.7) | 31.2 (13.0) | 29.3 (8.9) | 31.5 (11.6) | 28.4 (9.2) | 29.0 (6.3) |
| Trail B | 69.9 (29.9) | 71.6 (30.2) | 68.8 (31.2) | 74.1 (29.9) | 71.6 (25.9) | 69.8 (20.1) | 71.7 (24.2) | 71.7 (36.6) |
| Animals | 22.8 (5.2) | 23.3 (5.7) | 22.7 (5.5) | 23.1 (5.7) | 21.7 (5.5) | 22.1 (4.6) | 22.9 (5.5) | 24.0 (5.6) |
| Vegetables | 16.9 (3.9) | 16.7 (4.0) | 14.2 (3.7) | 14.5 (4.1) | 16.0 (3.5) | 16.4 (4.7) | 13.5 (3.2) | 13.4 (3.5) |

*Note:* CDR-SB is not included as values were 0 at baseline for all participants. Abbreviations: SCD: Subjective Cognitive Decline; APOE, apolipoprotein E; CDR-SB, Clinical Dementia Rating–Sum of Boxes; FAQ: Functional Activities Questionnaire.

Figure 1 and supplementary Table S1 summarize the estimated sloped differences between cognitive decline trajectories of SCD+ and SCD-groups. While SCD- and SCD+ groups did not differ in global cognition (as measured by CDR-SB) at baseline (*p* > 0.1), both groups showed significant decline in global cognition over time (*p* < .0001), and SCD+ individuals declined significantly faster than SCD-individuals (*β* = 0.023, *t* = 2.83, *p* = .004 for CDR-SB). In contrast, while both SCD groups showed decline in executive functioning over time (measured by Trail A and B, *p* < .0001), functional activity (measured by FAQ, *p* < .0001), and semantic fluency (measured by Animals and Vegetables, *p* < .0001), no statistically significant interaction (***Time from Baseline :* SCD**) differences were observed (*p* > 0.05). Finally, SCD+ participants showed marginally worse baseline performance in semantic fluency (Vegetables, *β* = 0.012, *t* = 2.00, *p* = .045) and executive functioning (Trail A, *β* = 0.012, *t* = 1.91, *p* = .054).

**Figure 1.**
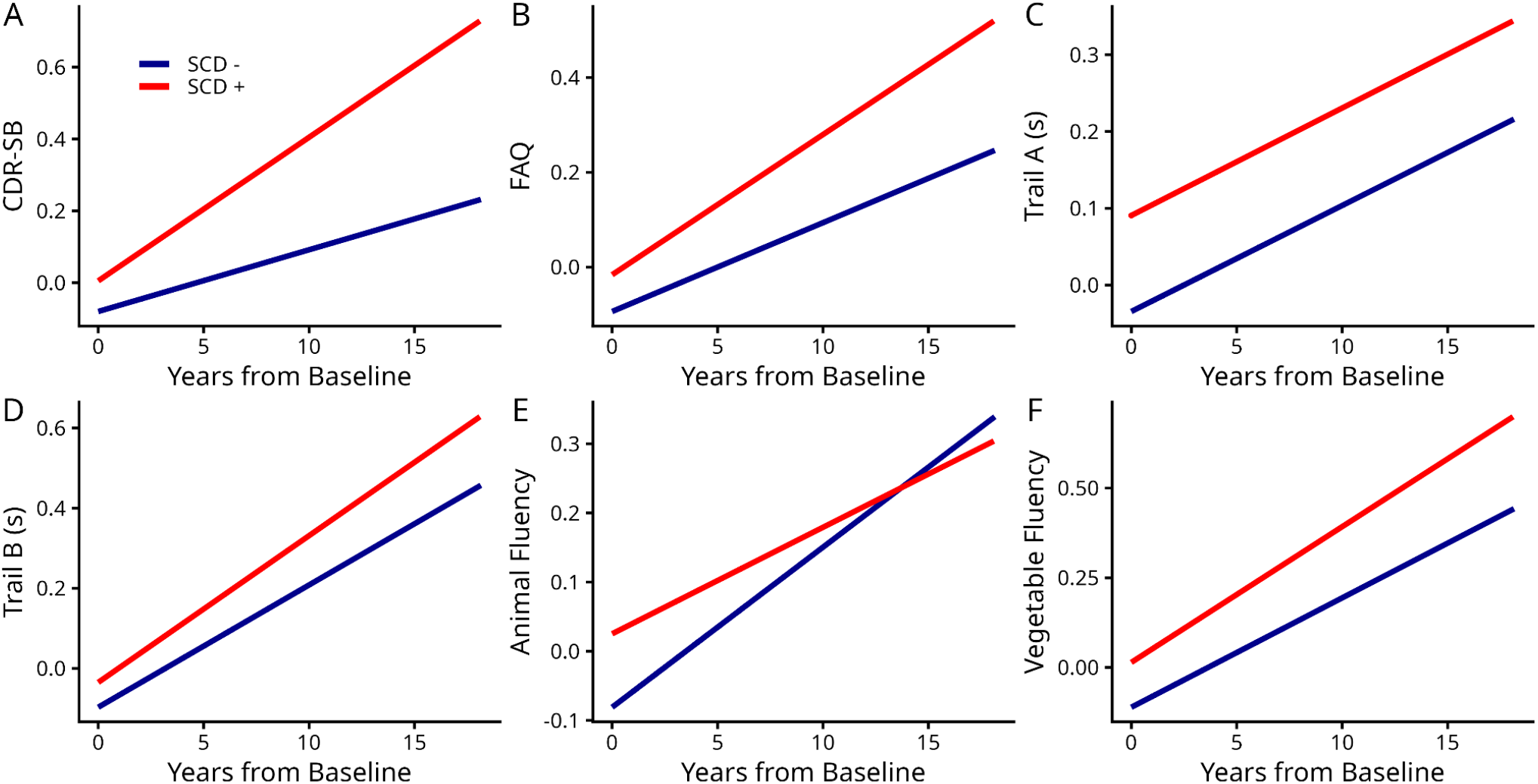
Cognitive decline trajectories across SCD status groups. Note: CDR-SB, Clinical Dementia Rating–Sum of Boxes. All groups declined over time for all cognitive measures and SCD+ exhibited steeper decline than SCD-in CDR-SB and Animal and Vegetable Fluency measures.

Figure 2 and Supplementary Table S2 summarize the estimated slope differences between cognitive decline trajectories of SCD and amyloid (A) classifications across different cognitive domains. All groups showed significant cognitive decline in global cognition (CDR-SB) and functional activity (FAQ) in the study follow-up duration (*p* < .0001). While A-SCD+ individuals did not experience faster decline in global cognition compared to A-SCD-individuals in CDR-SB and FAQ, A+SCD-individuals showed faster rates of cognitive decline (*β* = 0.003, *t* = 2.01, *p* = .04 for CDR-SB and *β* = 0.014, *t* = 1.98, *p* = .04 for FAQ) than A-SCD-. There was also a significant interaction between amyloid positivity and SCD status (*β* = 0.02, *t* = 2.80, *p* = .005 for CDR-SB and *β* = 0.030, *t* = 1.98, *p* = .04 for FAQ), indicating that A+SCD+ experienced exacerbated global and functional cognitive decline than all other groups. Furthermore, all groups showed declines in semantic fluency (*p* < .0001), with the A+SCD+ group experiencing marginally faster declines compared to the reference A-SCD-group (*β* = 0.15, *t* = 1.90, *p* = .05 for Animals). In contrast, while all groups showed significant decline in executive functioning (Trail A and B), no statistically significant group differences were observed in the trajectories of cognitive decline for Trail A, and only A+SCD-individuals showed significantly faster decline compared to A-SCD-individuals (*β* = 0.919, *t* = 3.84, *p* < .001). While the A+SCD+ group had a similar estimated slope to the A+SCD-, its slope difference did not reach statistical significance due to its smaller sample size.

**Figure 2.**
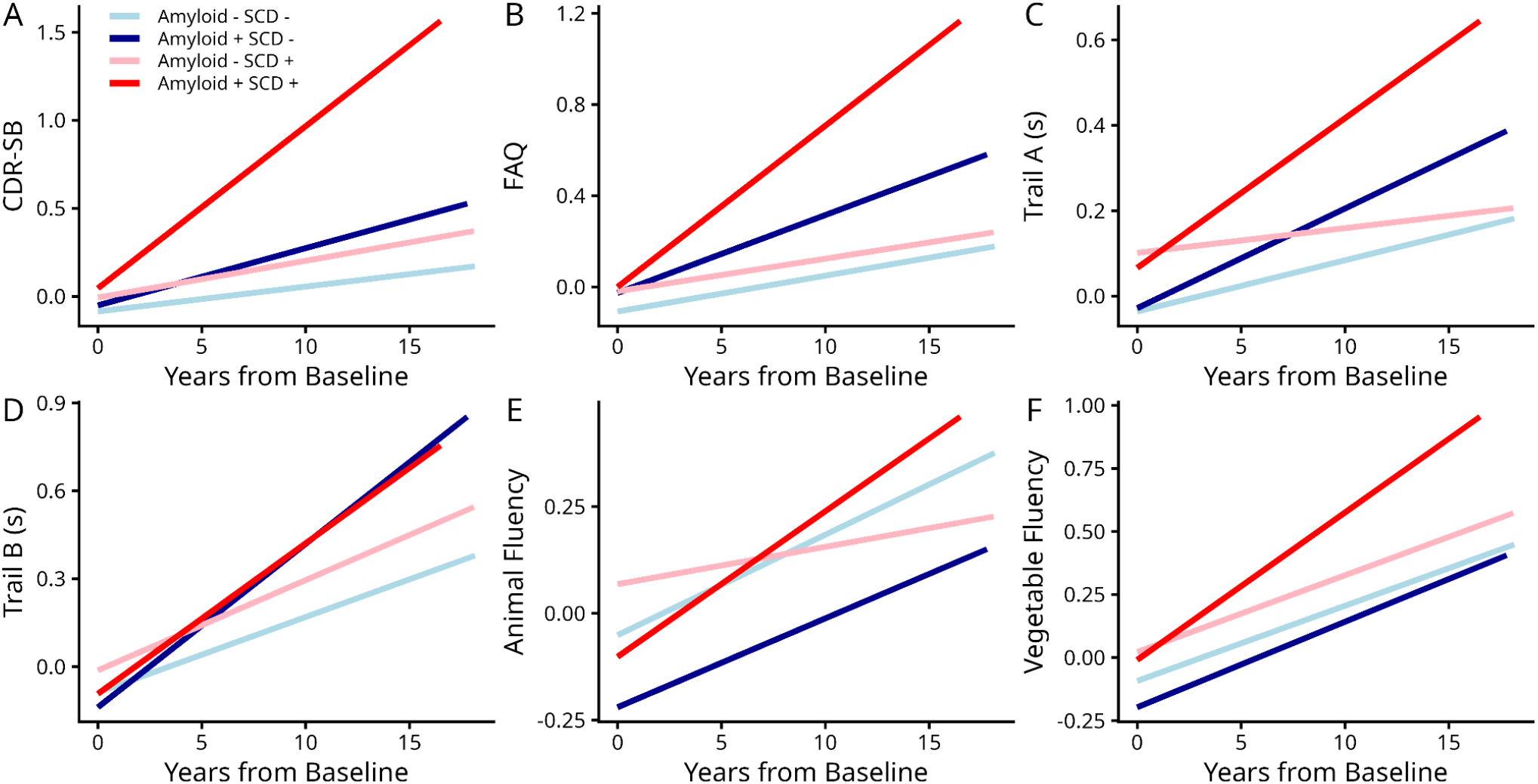
Cognitive decline trajectories across amyloid positivity and SCD status groups. Note: CDR-SB, Clinical Dementia Rating–Sum of Boxes; FAQ, Functional Activities Questionnaire.

Figure 3 and Supplementary Table S3 summarize the results of the sex differences in the estimated slopes of the cognitive decline trajectories of SCD and amyloid status groups across different cognitive domains. While all groups showed significant declines in global cognition (CDR-SB, *p* < .003), only A+SCD+ males showed significantly faster rates of decline compared to A-SCD-males (*β* = 0.09, *t* = 2.54, *p* = .01). In contrast, A+SCD+ males (*β* = 0.08, *t* = 3.13, *p* = .001) and A-SCD+ females (*β* = 0.05, *t* = 2.95, *p* = .003) showed significantly faster declines in functional activity (FAQ) compared to A-SCD-males. Both A-SCD-(*β* = 0.017, *t* = 2.82, *p* = .004) and A+SCD-(*β* = 0.035, *t* = 2.42, *p* = .01) males showed significantly faster declines compared to A-SCD-females, with a significant 4 way interaction (*β* = 0.064, *t* = 2.01, *p* = .04), indicating that the A+SCD+ males had the fastest rates of cognitive decline. With regards to semantic fluency (Animals and Vegetables), A+SCD+ females (*β* = 0.26, *t* = 2.66, *p* = .007 for animals and *β* = 0.28, *t* = 3.42, *p* = .0006 for vegetables), A+SCD-males (*β* = 0.24, *t* = 3.09, *p* = .002 for animals and *β* = 0.31, *t* = 4.96, *p* < .0001 for vegetables), and A-SCD+ males (*β* = 0.15, *t* = 2.10, *p* = .03 for vegetables) showed faster decline than A-SCD-females. Finally, there was a significant 4 way interaction for vegetables, suggesting that A+SCD+ females showed faster decline (*β* = 0.45, *t* = 3.28, *p* = .001) compared to A-SCD-females, while males overall started at worse baseline performance levels (*β* = 2.80, *t* = 11.47, *p* < .0001). There were no statistically significant group differences in executive functioning (Trail A and B).

**Figure 3.**
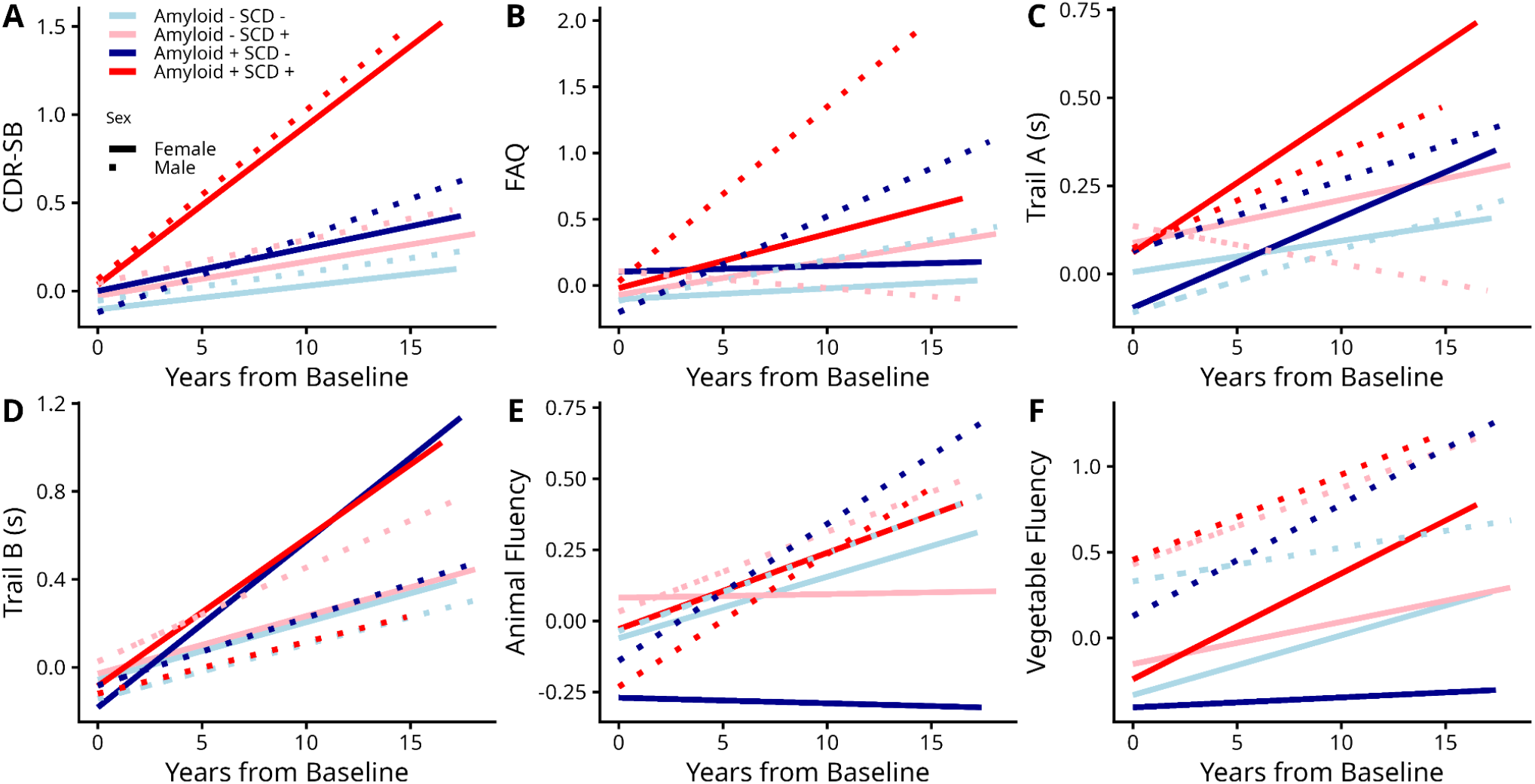
Cognitive decline trajectories across amyloid positivity, SCD status, and sex groups. CDR-SB, Clinical Dementia Rating–Sum of Boxes;

Figure 4 shows the percentage of participants that experience cognitive decline (drop in CDR-SB) in the duration of the study follow-ups (6.33 ± 4.2 years). Overall, 11.5% of the SCD+ group showed cognitive decline, compared to 6.7% of the SCD-group (*p* < .05). Within Amyloid and SCD categories, A-SCD-individuals had the lowest rate of decline (6%), followed by A-SCD+ (9%) and SCD-A+ (9.9%) groups, while the A+SCD+ had the highest rate of decline (18.6%, *p* < .005 contrasted against the A-SCD-group). Finally, A-SCD-females had the lowest rate of decline (5%), followed by A-SCD-males (7.7%), A-SCD+ (8.4%) and A+SCD-(8.9%) females, A-SCD+ (10.4%) and A+SCD-(11.4%) males, and A+SCD+ females (18.4%) and A+SCD+ males (19%) had the highest rates of decline, although none of the differences reached statistical significance due to the small subgroup sample sizes.

**Figure 4.**
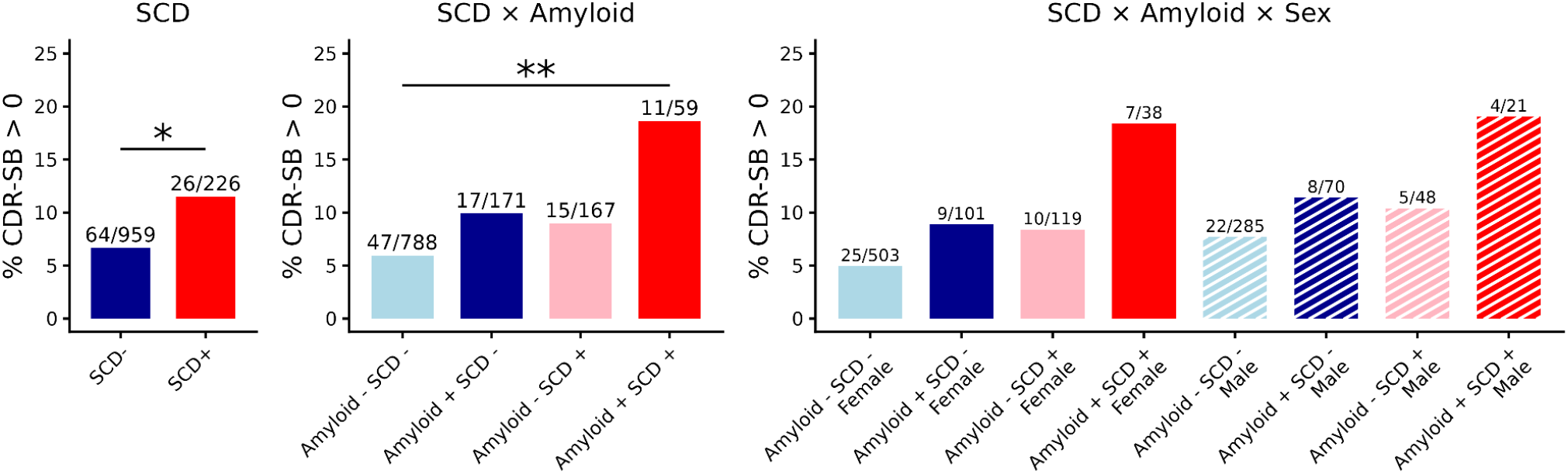
Differences in the rates of cognitive decline. Differences in the rates of cognitive decline based on increase in CDR-SB across groups between SCD (left), SCD and amyloid (middle), and SCD, amyloid, and sex (right) groups. Note: * FDR-corrected p < 0.05. ** FDR-corrected p < 0.005.

## Discussion

The current study examined the longitudinal trajectories of cognitive decline in cognitively intact aging individuals based on their amyloid positivity and SCD status, and further examined whether sex influenced these trajectories. Our findings showed significant differences in the rates of decline across groups over study duration, as well as domain specific differences in these rates of decline across SCD and amyloid groups. SCD+ individuals showed significantly faster decline in global cognition, but not executive functioning or functional activity. Furthermore, A+SCD+ individuals showed faster decline in global cognition and functional activity compared to all other groups, indicating an additive effect of SCD and amyloid positivity on global cognition and functional abilities. Amyloid positivity was also shown to confer significantly higher risk of decline in males compared to females. More specifically, the A+ male group showed significantly faster decline in functional activity compared to other groups.

While all groups showed significant cognitive decline over the duration of the study (mean follow up = 6.33 ± 4.2 years), significant differences emerged across SCD status, amyloid positivity, and sex. In line with previous research, SCD+ individuals had increased rates of decline compared to SCD-^4–6^, however, we also observed that this rate of decline was approximately 1.7 times higher in SCD+ vs SCD-(11.5% for SCD+ vs 6.7% for SCD-). This decline was observed in global cognition as assessed by the CDR-SB, a measure sensitive to early AD^37,38^. On the other hand, differences were not observed due to SCD status in executive functioning (as measured by processing speed) and functional status. One possible explanation for no group differences in these domains is that the processing speed measure may not have been complex or sensitive enough to detect subtle changes that may occur in the preclinical stages, particularly given that executive function, specifically processing speed, deficits are not typically the earliest indicators of AD^39^. Additionally, functional impairments generally manifest after the onset of cognitive symptoms^20,40^, which may explain the lack of observed differences in functional status due to SCD status.

Further within group comparisons pinpointed the highest rate of decline to the A+SCD+ group (18.6%), and this drastic increase was reflected in both females (18.4%) and males (19%). This finding is consistent with previous research indicating that amyloid positivity increases the rate of decline in people with SCD and that those with amyloid positivity are more likely to convert to AD than those without amyloid positivity^1–3,23^. In the current study, amyloid positivity was specifically related to increased declines in functional status and global cognition. Therefore, while SCD mainly impacted global cognition, amyloid positivity influenced global cognition, semantic fluency, and functional status, suggesting that the declines related to SCD and amyloid positivity may be domain specific or sensitive to only complex tasks (i.e., not processing speed as measured here). We also observed an additive effect of amyloid positivity and SCD status on semantic fluency within females only; those who were A+SCD+ exhibit increased declines in vegetable fluency compared to A-SCD-. This finding is important to note as language declines are also an early indicator of future cognitive deficits^41^ and conversion to AD in those with mild cognitive impairment^42^. Our prior work has shown that SCD+ females exhibit increased rates of decline compared to SCD+ males^6^. However, in the current study, we observed that amyloid positive males had increased declines in functional activity compared to other groups. Taken together, these findings suggest that while females may be more vulnerable to SCD-related decline than males^6^, amyloid positivity may heighten functional declines among males. Previous research has also observed that males with amyloid positivity have increased decline compared to females^43^.

Our findings have important implications for research cohort and clinical trial enrichment. While SCD and amyloid positivity alone conferred modest increases in risk of decline (9 and 9.9%, respectively compared to 6% in A-SCD-individuals), endorsing both conditions more than tripled the risk of decline (18.6%), suggesting that SCD evaluation is an effective strategy to pinpoint individuals that are likely to experience decline. This finding is particularly relevant as amyloid clearing treatments are most effective at the earliest disease stages^44,45^, where removal of amyloid would be more likely to prevent subsequent neurodegeneration and cognitive decline. Importantly, this increase in CDR-SB was similar across both males and females (19% and 18.4% respectively), indicating that the approach would be effective for both sexes. However, amyloid positivity differentially influenced domain-specific decline in males vs females. In females with SCD, it was associated with language decline, whereas in males it was linked to increased functional deficits irrespective of SCD status. This pattern of greater functional impairment in amyloid-positive males may partly help explain the emerging evidence that amyloid-targeting treatments may result in greater clinical benefit in males^34^.

Although prior work has investigated the differences in cognitive decline trajectories across amyloid and SCD status^22–26^, they often had limited sample sizes, short follow-ups, and did not examine the interaction with sex across in the longitudinal trajectories across these groups simultaneously. Our study was enabled by the large sample size of participants with amyloid biomarkers, SCD classification, and longitudinal cognitive assessments provided by the NACC (1185 participants, 7443 longitudinal assessments, 6.33 ± 4.2 years of follow-up). Importantly, all groups had similar follow-up durations, preventing attrition related biases in the observed group differences and allowing us to robustly model the potential difference across groups of interest.

There are certain limitations within the current study. First, while our sample allowed for examination of the impact of amyloid status on the cognitive decline trajectories across sexes in those with SCD, the limited sample sizes of individuals who also had tau biomarkers or MRI prevented us from examining ATN-V differentiated trajectories. This limited sample likely reflects that individuals with greater pathological burden (tau, neurodegeneration, and vascular pathology) are likely to exhibit cognitive declin^20,46,47^. In AD, amyloid accumulation is predicted to occur earlier in the disease process^20^, with tau pathology and neurodegeneration more closely linked to cognitive symptoms. Thus, our focus on cognitively normal individuals (CDR-SB = 0) may have limited the inclusion of participants with more advanced pathology. Future studies within larger samples are warranted to assess the SCD trajectories across ATN-V groups and their potential sex differences.

Overall, these findings indicate that SCD and amyloid independently and jointly influence cognitive decline and functional status. Importantly, these effects vary across cognitive domains and functional abilities between males and females. Together, these findings highlight the heterogeneity of early cognitive decline and underscore the importance of considering both biological and sex-related factors when characterizing risk trajectories. Future research should explore how amyloid interacts with other biomarkers such as tau, neurodegeneration, and vascular pathology in SCD to improve our understanding of the full trajectory of change in SCD.

## Data Availability

All data produced in the present work are contained in the manuscript

## Acknowledgments

The NACC database is funded by NIA/NIH Grant U24 AG072122. NACC data are contributed by the NIAfunded ADRCs: P30 AG062429 (PI James Brewer, MD, PhD), P30 AG066468 (PI Oscar Lopez, MD), P30 AG062421 (PI Bradley Hyman, MD, PhD), P30 AG066509 (PI Thomas Grabowski, MD), P30 AG066514 (PI Mary Sano, PhD), P30 AG066530 (PI Helena Chui, MD), P30 AG066507 (PI Marilyn Albert, PhD), P30 AG066444 (PI David Holtzman, MD), P30 AG066518 (PI Lisa Silbert, MD, MCR), P30 AG066512 (PI Thomas Wisniewski, MD), P30 AG066462 (PI Scott Small, MD), P30 AG072979 (PI David Wolk, MD), P30 AG072972 (PI Charles DeCarli, MD), P30 AG072976 (PI Andrew Saykin, PsyD), P30 AG072975 (PI Julie A. Schneider, MD, MS), P30 AG072978 (PI Ann McKee, MD), P30 AG072977 (PI Robert Vassar, PhD), P30 AG066519 (PI Frank LaFerla, PhD), P30 AG062677 (PI Ronald Petersen, MD, PhD), P30 AG079280 (PI Jessica Langbaum, PhD), P30 AG062422 (PI Gil Rabinovici, MD), P30 AG066511 (PI Allan Levey, MD, PhD), P30 AG072946 (PI Linda Van Eldik, PhD), P30 AG062715 (PI Sanjay Asthana, MD, FRCP), P30 AG072973 (PI Russell Swerdlow, MD), P30 AG066506 (PI Glenn Smith, PhD, ABPP), P30 AG066508 (PI Stephen Strittmatter, MD, PhD), P30 AG066515 (PI Victor Henderson, MD, MS), P30 AG072947 (PI Suzanne Craft, PhD), P30 AG072931 (PI Henry Paulson, MD, PhD), P30 AG066546 (PI Sudha Seshadri, MD), P30 AG086401 (PI Erik Roberson, MD, PhD), P30 AG086404 (PI Gary Rosenberg, MD), P20 AG068082 (PI Angela Jefferson, PhD), P30 AG072958 (PI Heather Whitson, MD), P30 AG072959 (PI James Leverenz, MD).

## Consent Statement

Written informed consent was obtained from participants or their study partner.

## Availability of data and materials

The data utilized in this study were also sourced from the National Alzheimer’s Coordinating Center (NACC) database (https://naccdata.org/), specifically drawing from the NACC Uniform Data Set (UDS) and MRI Data Set (Beekly et al., 2004; Besser, Kukull, Knopman, et al., 2018; Besser, Kukull, Teylan, et al., 2018).

